# Exploring options for reprocessing of N95 Filtering Facepiece Respirators (N95-FFRs) amidst COVID-19 pandemic: a systematic review

**DOI:** 10.1101/2020.09.01.20179879

**Authors:** Diptanu Paul, Ayush Gupta, Anand Kumar Maurya

## Abstract

**Background:** There is global shortage of Personal Protective Equipment due to COVID-19 pandemic. N95 Filtering Facepiece Respirators (N95-FFRs) provide respiratory protection against respiratory pathogens including SARS-COV-2. There is scant literature on reprocessing methods which can enable reuse of N95-FFRs.

**Aim:** We conducted this study to evaluate research done, prior to COVID-19 pandemic, on various decontamination methods for reprocessing of N95-FFRs.

**Methods:** We searched 5 electronic databases (Pubmed, Google Scholar, Crossref, Ovid, ScienceDirect) and 1 Grey literature database (OpenGrey). We included original studies, published prior to year 2020, which had evaluated any decontamination method on FFRs. Studies had evaluated a reprocessing method against parameters namely physical changes, user acceptability, respirator fit, filter efficiency, microbicidal efficacy and presence of chemical residues post-reprocessing.

**Findings and Conclusions:** Overall, we found 7887 records amongst which 17 original research articles were finally included for qualitative analysis. Overall, 21 different types of decontamination or reprocessing methods for N95-FFRs were evaluated. Most commonly evaluated method for reprocessing of FFRs was Ultraviolet (Type-C) irradiation (UVGI) which was evaluated in 13/17 (76%) studies.

We found published literature is scant on this topic despite warning signs of pandemic of a respiratory illness over the years. Promising technologies requiring expeditious evaluation are UVGI, Microwave generated steam (MGS) and Hydrogen peroxide vapor (HPV). Global presence of technologies, which have been given Emergency use authorisation for N95-FFR reprocessing, is extremely limited. Reprocessing of N95-FFRs by MGS should be considered for emergency implementation in resource limited settings to tackle shortage of N95-FFRs.

**Systematic Review Identifier:** PROSPERO, PROSPERO ID: CRD42020189684, (https://www.crd.york.ac.uk/prospero/display_record.php?ID=CRD42020189684)

## Introduction

Global pandemic of Corona Virus Disease of 2019 (COVID-19) has led to over 10 million cases and half-a-million deaths worldwide and still counting[1]. It is caused by a novel Corona virus (nCOV), a member of family *Coronaviridae*, now renamed as SARS-COV-2[2]. Transmission of this virus occurs through direct, contact and airborne routes[3]. Consequently, healthcare workers (HCWs) requires a full set of personal protective equipment (PPE) including gowns, gloves, facemasks, face-shields or goggles and respirators for their protection during patient care[4]. This has created an unprecedented demand for PPEs leading to their global shortage forcing administrative authorities to relook the recommendations of PPE usage in a whole new light[5]. Previously, focus of PPE use strategy was not to share them between patients[6] however, due to this unprecedented crisis, it has radically shifted to optimizing the use of PPEs, their extended use and limited reuse[4,5,7]. Respiratory protection is one of the fundamental rights of any employee in workplace. In healthcare settings, HCWs need to be protected against bioaerosols at all costs, which at minimum, is offered by use of N95 Filtering Facepiece Respirator (N95-FFR) which removes > 95% particles of around 300 nm[6]. They are single use devices ought to be discarded after use to avoid cross contamination[8].

Shortage of FFRs is not new, pangs of which were first felt during 2003 SARS outbreak[9]. It has been predicted for an impending influenza pandemic consequent to which U.S. Strategic National Stockpile had plans for providing 100 million N95-FFRs nationally, but it was deemed insufficient in event of a longer pandemic[9-11]. Hence, in 2006, Institute of Medicine (IOM) constituted a committee to address reusability of facemasks. Reuse of an FFR was defined as repeatedly donning and doffing of respirator by the same wearer, with or without undergoing reprocessing in between, till it is discarded. The committee recommended reuse of respirators in the event of acute shortage provided they are not obviously damaged or soiled[11]. However, committee specified that no method exists currently for reprocessing of N95-FFRs and identified it as a research priority[11]. Consequently, various research groups began their quest to search a reprocessing method which is efficacious against respiratory pathogens, is safe for human use and maintains the integrity of various components of the respirator. Even after a decade of research, prior to COVID-19 pandemic, no method has been recommended for reprocessing of N95-FFRs. Hence, we conducted this systematic review to determine the status of research done, prior to COVID-19 pandemic, to identify technologies which can be utilized for reprocessing of N95-FFRs in present situation and can be explored in near future to tackle the global crisis of respirator shortage.

## Methods

We report this systematic review (PROSPERO ID: CRD42020189684) in accordance with the Preferred Reporting Items for Systematic Reviews and Meta-Analyses (PRISMA) guidelines[12] and checklist is provided in S1 Table.

### Search strategy

We searched five databases – Pubmed, Google Scholar, Crossref, Ovid and ScienceDirect in May 2020. Grey literature was searched using OpenGrey repository. Search strategies employing combinations of various keywords is provided in S2 Table. Searches in Google Scholar and Crossref were done using Publish or Perish 7 software (Harzing, A.W. 2007) to limit article hits and sort relevant ones. Additionally, we manually searched the back references of included studies and relevant review articles on the topic to identify further eligible studies. Articles in languages other than English were considered only when their abstracts were available in English.

### Eligibility criteria

Original research articles in any language, which evaluated a single or multiple decontamination or reprocessing methods on N95-FFRs were eligible for analysis in this study. Exclusion criteria were (i) Abstracts, posters, review articles, book chapters, letters, guidelines, point of views (ii) articles published in year 2020 and (iii) involving reprocessing or decontamination of other types of masks or respirators such as Gauze, Cloth, Spun-lace, Elastomeric and Powered-air-purifying, only.

### Data Extraction

After searching all databases, we exported data in Microsoft® Excel and removed duplicates. Two reviewers (DP & AG) screened titles to remove clearly irrelevant studies. All three reviewers (AG, DP, AKM) independently screened the abstracts and full text of remaining articles to determine final eligibility and resolved any discrepancies through discussion and consensus. After included studies were finalized, data on various variables such as reprocessing method exposure variables, number, type and replicates of FFR models, parameters which were evaluated and final results was entered in Microsoft® Excel independently by all three reviewers. Extracted data was checked and analysed by one reviewer (AG) and disagreements were resolved prior to final analysis.

### Quality Assessment

To assess methodological quality and risk bias of studies, a self-developed tool was designed on the basis of STROBE statement[13] due to unavailability of a validated quality assessment tool for such studies. Two authors (AKM and DP) independently assessed the methodological quality and risk bias as per tool. The scheme of scoring and grading of studies is given in S3 Table along with the final quality assessment results. Inter-author concordance on grading of studies was evaluated by third author (AG). Final quality assessment results for included studies, as shown in S3 Table, were prepared by resolving inter-author disagreements by discussion and building consensus.

## Results

### Search Results

Our search strategy identified 7887 records of which 17 original research articles fit inclusion criteria for qualitative analysis, methodology of the same has been described in Fig 1. No records were found in OpenGrey database using search strategy.

**Fig 1.**
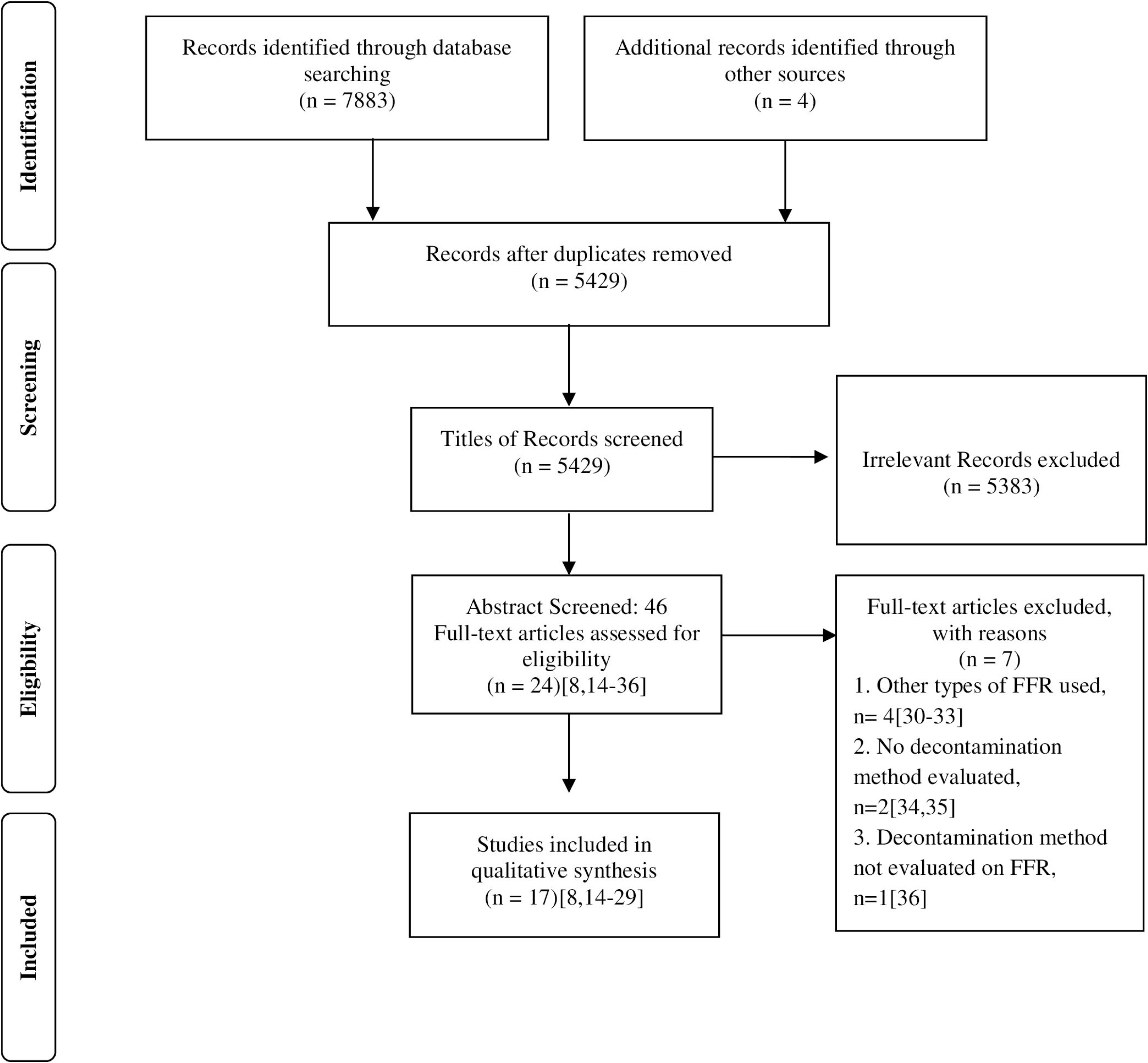
Summary of search, selection and inclusion process. Legend: Abbreviations: FFR: Filtering Facepiece Respirator, n: Number

### Quality Assessment

Of 17 studies, 14 were graded as high quality and 3 as moderate quality (S3 Table). Inter-author agreement in grading of studies was 88% (15/17). Overall agreement in quality assessment scores was 64% (11/17).

### Study Characteristics

Amongst 17 included studies, 15 were conducted in U.S.[8,14-27] and 2 in Taiwan[28,29]. Ten out of 15 studies were conducted by research groups from NIOSH as the principal investigator[8,14,16,17,21-26], 4 by researchers at Applied Research Associates (ARA) in collaboration with Air Force Research Laboratory at Tyndall Air Force Base, Panama City[18-20,27] and in 1 study, principal investigators were from University of Nebraska (UoN)[15]. Three studies were an outcome of collaboration between NIOSH, ARA & UoN in various combinations[14,15,18]. Two studies from Taiwan were conducted by same researchers at Department of Occupational Safety and Health, Chung Shan Medical University[28,29]. First study evaluating reprocessing methods for FFRs was published in 2007[22] and last study in 2018[29].

### Decontamination/ Reprocessing methods

Overall, 21 different types of decontamination or reprocessing methods for N95-FFRs were evaluated in included studies against various parameters namely physical changes, user acceptability, respirator fit, filter efficiency, microbicidal efficacy and presence of chemical residues post-reprocessing. Number of studies conducted for each reprocessing method, on these parameters are given in Fig 2. Overall, these studies evaluated 9 Physical (Energetic) reprocessing methods namely Ultraviolet (UV-C) Irradiation (UVGI)[8,14-16,19,20,23-25,27,29], UV-A[29], UV-B[27], Moist heat delivered using Microwave generated Steam (MGS)[14,15,20,22,23,26], Lab Incubator (MHI)[14,15,20,22,23] and Autoclave (MHA)[22,28,29], Dry heat delivered by Microwave (MGI)[16,22], Hot Air Oven (DHO)[22] and Traditional Electric Rice Cooker (TERC)[28,29]; 3 Gaseous chemical decontamination methods namely Hydrogen Peroxide Gas Plasma (HPGP)[14,16,22,27], Hydrogen Peroxide Vapor (HPV)[14] and Ethylene Oxide (EO)[14,16,22,27]; 6 Liquid chemical decontamination methods namely Bleach[14,16,22,25-29], Hydrogen Peroxide (LHP)[14,22], Alcohols[22,28,29], Mixed Oxidants[27], Dimethyl dioxirane[27] and Soap & water[22]; and in one study[18], wipes of Bleach (0.9%), Benzalkonium chloride and Inert substance for surface decontamination of N95-FFRs. Fourteen (14) studies[14-18,20-29] did comparative evaluation of multiple methods for reprocessing of FFRs whereas in 3 studies only 1 method was evaluated, which was UVGI in all[8,19,24]. In 12 studies[14-16,16-23,25,27], intact respirators were exposed to the decontamination method whereas in 5, cut pieces of facepiece portion were exposed[8,24,26,28,29]. Furthermore, in one study,^8^ pieces of straps were also exposed separately to UVGI. In 4 studies, FFRs underwent multiple cycles (3 in all studies) of decontamination for reprocessing[14,17,18,23].

**Fig 2.**
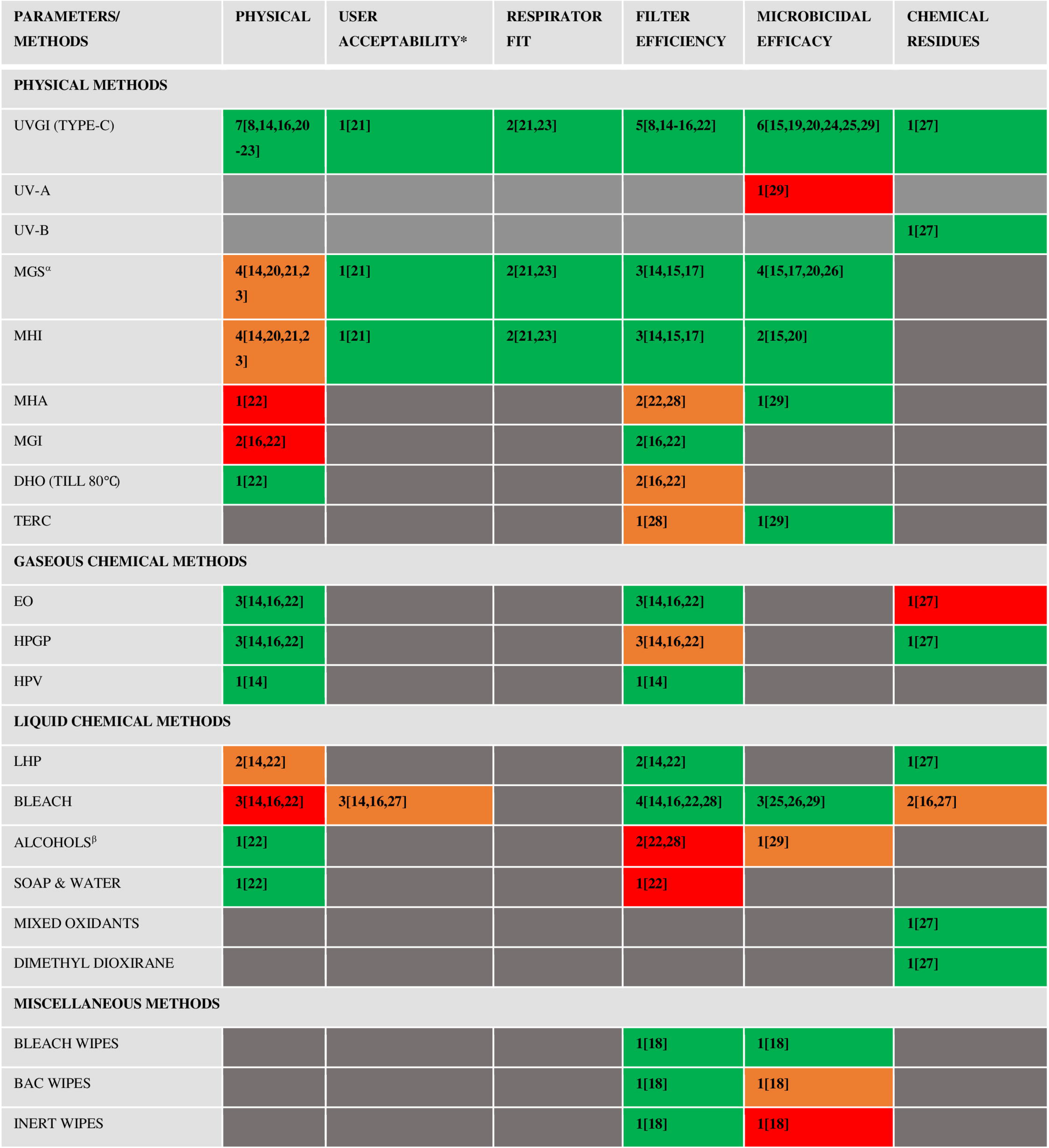
Summary of studies [Total Number, n^reference^] conducted on various parameters related to reprocessing of N95 Filtering Facepiece Respirators (FFRs), plotted against the reprocessing method. Coloured cells represent cumulative results of these studies (See Legend Below) Legend: Numbers in each coloured cells represent total number of studies conducted on a reprocessing method: parameter combination. Numbers in Superscript denote the reference number of studies, **Green Cells:** Evidence shows no negative effect of the reprocessing method on the evaluated parameter **Red Cells:** Evidence shows a negative effect of the reprocessing method on the evaluated parameter **Orange Cells:** Evidence shows an effect which is either in conflict in different studies or requires careful consideration **Grey Cells:** No study done on the reprocessing method: parameter combination * User Acceptability is a composite parameter including odour, wear comfort & donning ease. References 14,16,27 only evaluated odour α- Fisher *et al* 2011^17^ used Commercial steam bags for generation of steam, other studies used a water reservoir β- Ethanol (70%)^28,29^ and Isopropyl alcohol (70%^28^ and 100%^22^) were used Abbreviations: **UVGI:** Ultraviolet Irradiation (Type-C, 254 nm), **MGS:** Microwave Generated Steam, **MHI:** Moist heat Incubation in Lab Incubator, **MHA:** Moist Heat in Autoclave, **DHO**: Dry Heat in Oven (Till 80°), **TERC:** Traditional Electric Rice Cooker, **EO**: Ethylene Oxide, **HPGP**: Hydrogen Peroxide Gas Plasma, **HPV**: Hydrogen Peroxide Vapor, **LHP**: Liquid Hydrogen Peroxide, **BAC:** Benzalkonium Chloride Note: The summary is only indicative of the collective results of various studies done (prior to 2020) to evaluate effect of reprocessing method on a particular parameter. It doesn’t attempt to endorse or refute any method as the authors strongly believe that there is insufficient data to reach any conclusion.

### Respirator Models

In 10 of 17 studies, the identities of N95-FFR models used was disclosed[8,15,17,19,21,23-25,28,29], details of which against the reprocessing method and parameters evaluated are given in S4 Table. Overall, 22 different models of N95-FFRs were disclosed in 10 studies, 18 of which are approved as surgical respirators by FDA, whereas 4 are Particulate respirators. All respirators used in these studies, irrespective of whether identities were disclosed or not, were NIOSH approved. 3M1860 and 3M1870, both surgical respirators, were the most commonly used N95-FFRs, being used in seven[8,15,19,21,22,23,26] and six[15,17,19,21,23,26] studies, respectively. They were tested against three reprocessing methods i.e. UVGI, MGS and MHI, where identity was disclosed. 3M8210, a particulate respirator, was exposed to 7 different reprocessing methods. Furthermore, in 2 studies, P100 respirators were also evaluated but in both identities were not disclosed[16,22].

### Decontamination Methods

#### A. Physical (Energetic) Methods

##### i. Ultra-Violet Irradiation (UVGI)

Thirteen studies[8,14-16,19-25,27,29] evaluated exposure to UV-C (254 nm) as a reprocessing method for FFRs, as shown in Fig 2. All 22 known models of N95-FFRs were reprocessed using UV-C in at least one study (S4 Table). Furthermore, one study each also examined the microbiological efficacy of UV-A[29] and presence of chemical residues after using UV-B[27]. Exposure variables of UVGI on N95-FFRs and summary of results are provided in Table 1. Different parameters evaluated against UVGI are detailed in Fig 2. Since, UV-C has been the most commonly evaluated method for reprocessing of N95-FFRs, it will be discussed in detail.

**Table 1:** Summary of characteristics of studies using Ultraviolet Irradiation (UVGI) as a reprocessing method for N95-FFRs.

| **Authors** |  | **Variables of UVGI Irradiation** |  |  |  |  | **Variables of FFRs** |  |  | **Results** |  |
| --- | --- | --- | --- | --- | --- | --- | --- | --- | --- | --- | --- |
| **(Year)** | **Type** | **Irradiance**<br>**(mW/cm<sup>2</sup>)** | **Duration** | **Dose**<br>**(J/cm<sup>2</sup>)** | **Sides**<br>**Exposed**<br>**to UVGI** | **No. of**<br>**Cycle** | **Total**<br>**no. of**<br>**Models**<br>**used** | **Part of**<br>**FFR**<br>**exposed**<br>**to UVGI** | **Replicates** | **Parameters**<br>**Assessed** | **Summary of Results** |
| Bergman<br>*et al*[14]<br>(2010) | C | 1.8 | 45 m | - | Outer<br>(Convex) | 3 | 6 | Intact | 3 | Physical<br>Changes<br>Odour<br>Filter<br>Efficiency | No observable physical changes on FFRs<br>No comment on odour<br>Expected levels of Filter Aerosol penetration (<5%) & filter airflow resistance |
| Lore *et al*[15]<br>(2012) | C | 1.6-2.2 | 15 m | 1.8 | Outer<br>(Convex) | 1 | 2 | Intact | 9 | Filter<br>Efficiency<br>Microbicidal<br>Efficacy | No significant degradation of filter performance<br>>4 log<sup>10</sup> TCID<sub>50</sub>/ml reduction of H5N1 Avian Influenza virus |
| Viscusi *et al*[16]<br>(2009) | C | 0.18-0.2 | 30 m | 0.17-0.18 | Each side | 1 | 9 | Intact | 3 | Physical Changes<br>Filter Efficiency | No observable physical changes on FFRs<br>Didn't affect Filter efficiency |
| Lindsley *et al*[8]<br>(2015) | C |  |  | 120, 240, 470, 950 (For mask layers);<br>590, 1180, 2360 (For straps, each side) | NA | 1 | 4 | Facepiece<br>Coupons<br>and Straps | 4 | Structural Integrity<br>Filter Efficiency | Strengths of respirator materials was substantially reduced (in some cases>90%)<br>Slight increase in particle penetration but no effect on airflow resistance |
| Mills *et al*[19]<br>(2018) | C | 17 | 60-70 s | 1 | Outer (Convex) | 1 | 15 | Intact | 3 | Microbicidal Efficacy | $\geq 3 \log_{10}$ TCID<sub>50</sub>/ml reduction in Influenza virus (H1N1) viability on 12/15 FFR models and straps from 7/15 FFR models |
| Heimbuch | C | 1.6- | 15 m | 1.8 | Outer | 1 | 6 | Intact | 3 | Physical | No observable physical changes on |
| *et al*[20]<br>(2011) |  | 2.2 |  |  | (Convex) |  |  |  |  | Changes<br>Microbicidal<br>Efficacy | FFRs<br>>4 log<sub>10</sub> TCID<sub>50</sub>/ml reduction of<br>Influenza virus (H1N1) |
| Viscusi *et al*[21]<br>(2011) | C | 1.8 | 30 m | - | Each<br>side | 3 | 6 | Intact | 2 | Physical<br>Changes<br>User<br>Acceptability<br>Respirator Fit | No observable physical changes on<br>FFR<br>No clinically meaningful reduction<br>in respirator fit, increase in odour,<br>increase in discomfort or increased<br>difficulty in donning |
| Viscusi *et al*[22]<br>(2007) | C | 0.24 | 15/ 240 m | - | Each<br>side | 1 | 2 | Intact | 4 | Physical<br>Changes<br>Filter<br>Efficiency | No observable physical changes on<br>FFRs<br>Not significantly affected by both<br>time durations on both types of<br>FFRs (N95 and P100) |
| Bergman<br>*et al*[23] | C | 1.8 | 15 m | - | Outer<br>(Convex) | 3 | 3 | Intact | 2 | Physical<br>Changes | No observable physical changes on<br>FFRs |
| (2011) |  |  |  |  |  |  |  |  |  | Respirator Fit | No significant changes in Respirator fit |
| Fisher *et al*[24]<br>(2010) | C | 2.5 | 1, 2, 4, 10 m on 3M 8210,1870; 10 m on Cardinal N95-ML | 0.03, 0.1 and 0.3 on Wilson, 3M 1860 and KC | Each side | 1 | 6 | Facepiece Coupons | 3 | IFM specific dose for Microbicidal Efficacy | Log Reduction of MS2 Coliphage is a function of FFR model specific IFM UV-C dose |
| Lin *et al*[29]<br>(2018) | C | 18.9 | 1, 2,5, 10, 20 m | - | NA | 1 | 1 | Cut pieces | 3 | Microbicidal Efficacy | 99-100% biocidal efficacy against *Bacillus subtilis* spores |
| Vo *et al*[25]<br>(2009) | C | 0.4 | 1, 2, 3, 4, 5 hr | 1.44, 2.88, 4.32, 5.76, 7.2 | One side | 1 | 1 | Intact | 3 | Microbicidal Efficacy | 3 log reduction of MS2 Coliphage at dose of 4.32 J/cm<sup>2</sup> and complete removal at dose of ≥7.2 J/cm<sup>2</sup> |
| Salter *et al*[27] | C | 3.4 | 1 hour | 27 | NA | 1 | 6 | Coupons, straps, | 3 | Presence of Toxic | No toxic residues post-exposure |
| (2010) |  |  |  |  |  |  |  | Nose cushion,<br>Nose pieces |  | Chemical residues<br>Post-exposure |  |
| Lin *et al*[29]<br>(2018) | A | 31.2 | 1, 2,5, 10, 20 m | - | Each side | 1 | 1 | Cut pieces | 3 | Microbicial Efficacy | Poor Microbicial efficacy against *Bacillus subtilis* spores |
| Salter *et al*[27]<br>(2010) | B | 4 | 1 hr | - | NA | 1 | 6 | Coupons, straps,<br>nose cushion,<br>Nose pieces | 3 | Presence of Toxic Chemical residues<br>Post-exposure | No toxic residues post-exposure |

**ABBREVIATIONS:** **mW/cm^2^**: milli watt per square centimetre, **J/cm^2^**: Joules per square centimetre **m**: Minute, **NA**: Not Applicable, **FFR**: Filtering Facepiece Respirator, **TCID**: Tissue Culture Infectious Dose, **s**: seconds **IFM**: Internal Filtering Media, **hr**: Hour

##### ii. Moist Heat

Delivering moist heat to FFRs has been evaluated in 10 studies[14,15,17,18,21-23,26,28,29]. Modalities of exposure involved exposing FFRs to steam created in a microwave (MGS), either by using water reservoir[14,15,20,21,23,26] or commercial steam bags[17]; in a lab incubator with a water reservoir heated at 60-70°C (MHI)[14,15,18,21,23] and by autoclaving at 121°C (MHA)[22,28,29]. Parameters evaluated for these treatments are given in Fig 2 and the exposure variables and results of individual studies are described in Table 2. Known FFR models which underwent reprocessing by both MGS and MHI were 3M1860, 3M1870, 3M8000 and 3M8210, whereas, for MHA only known FFR model was 3M8210.

**Table 2:**
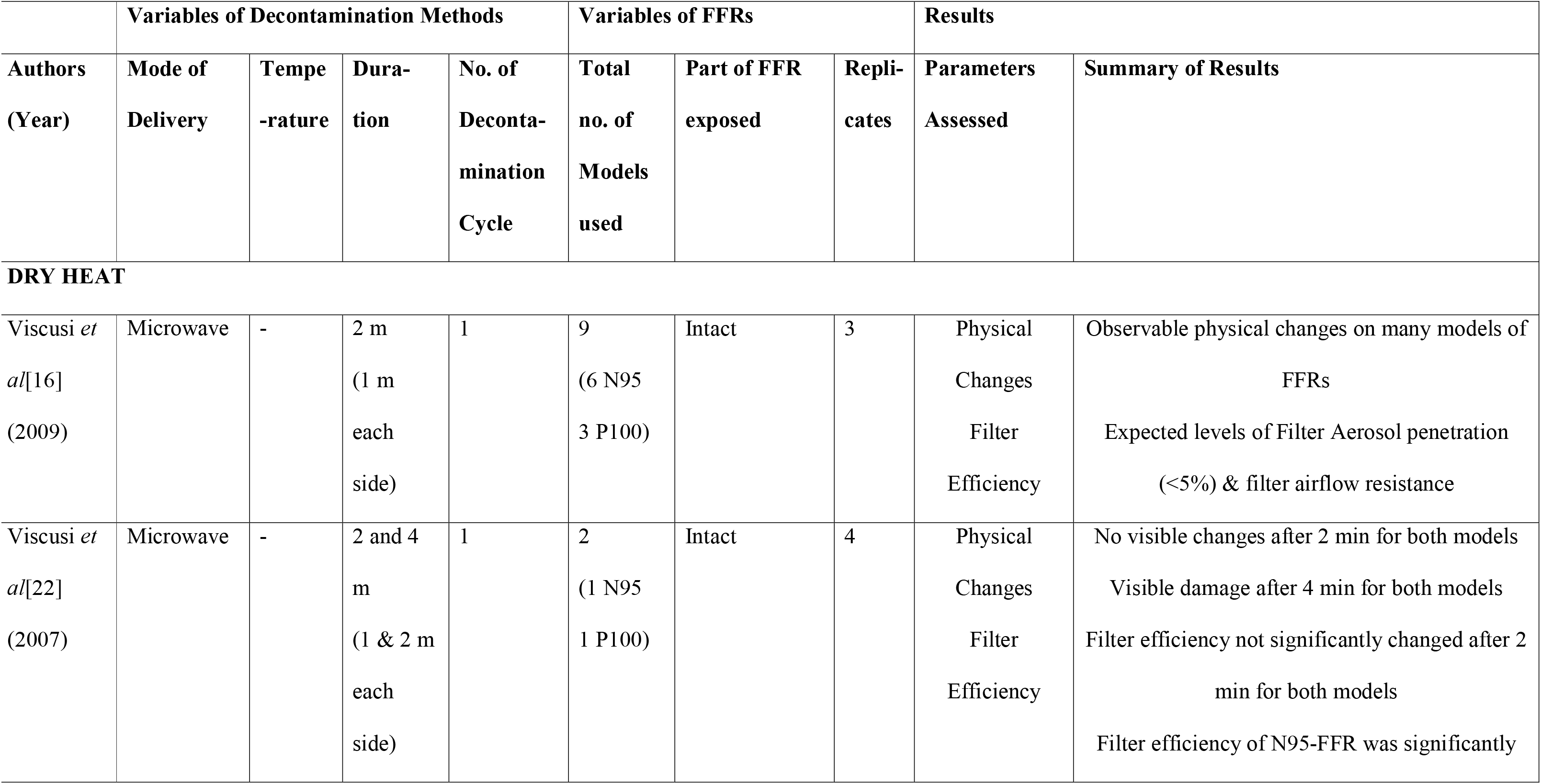
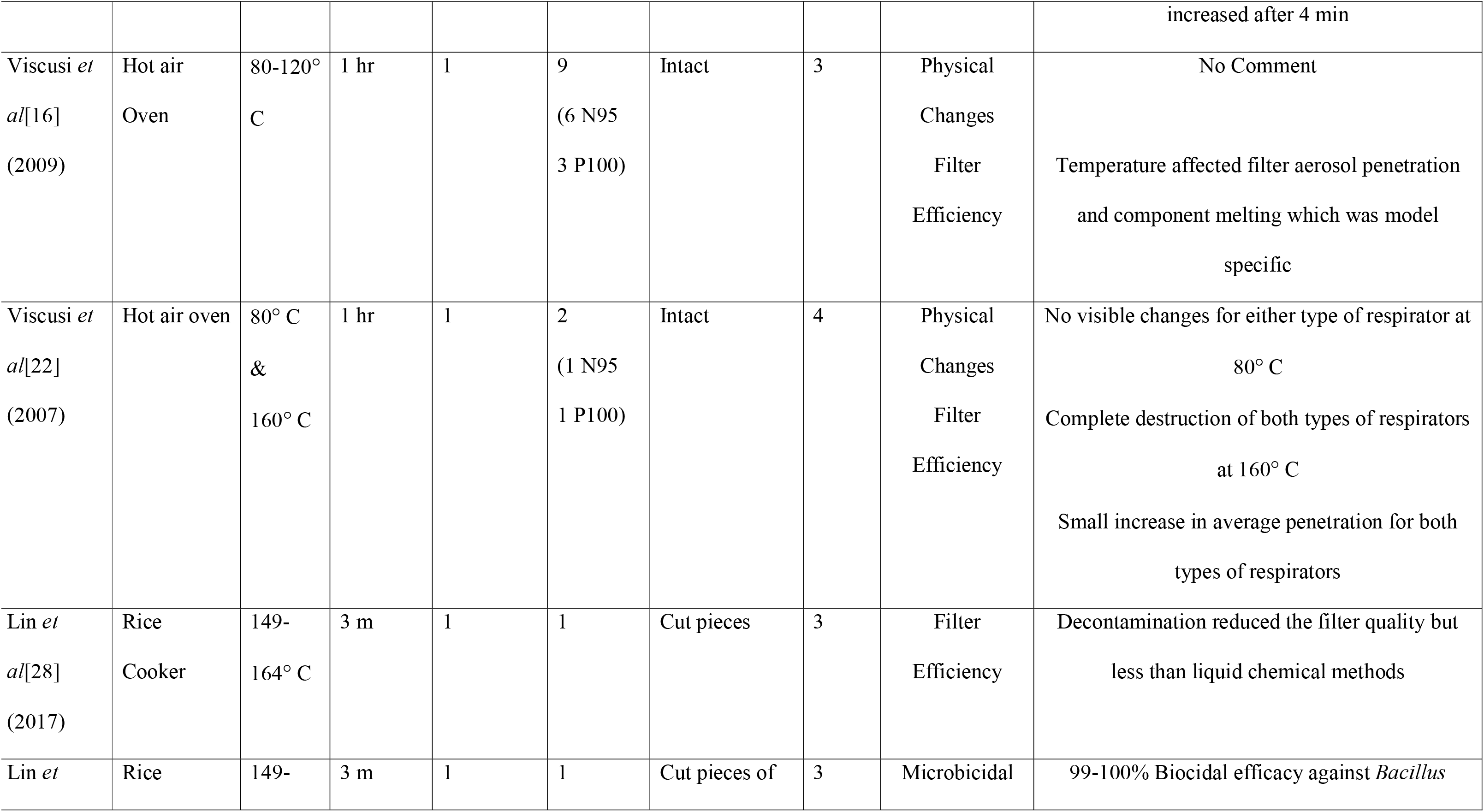
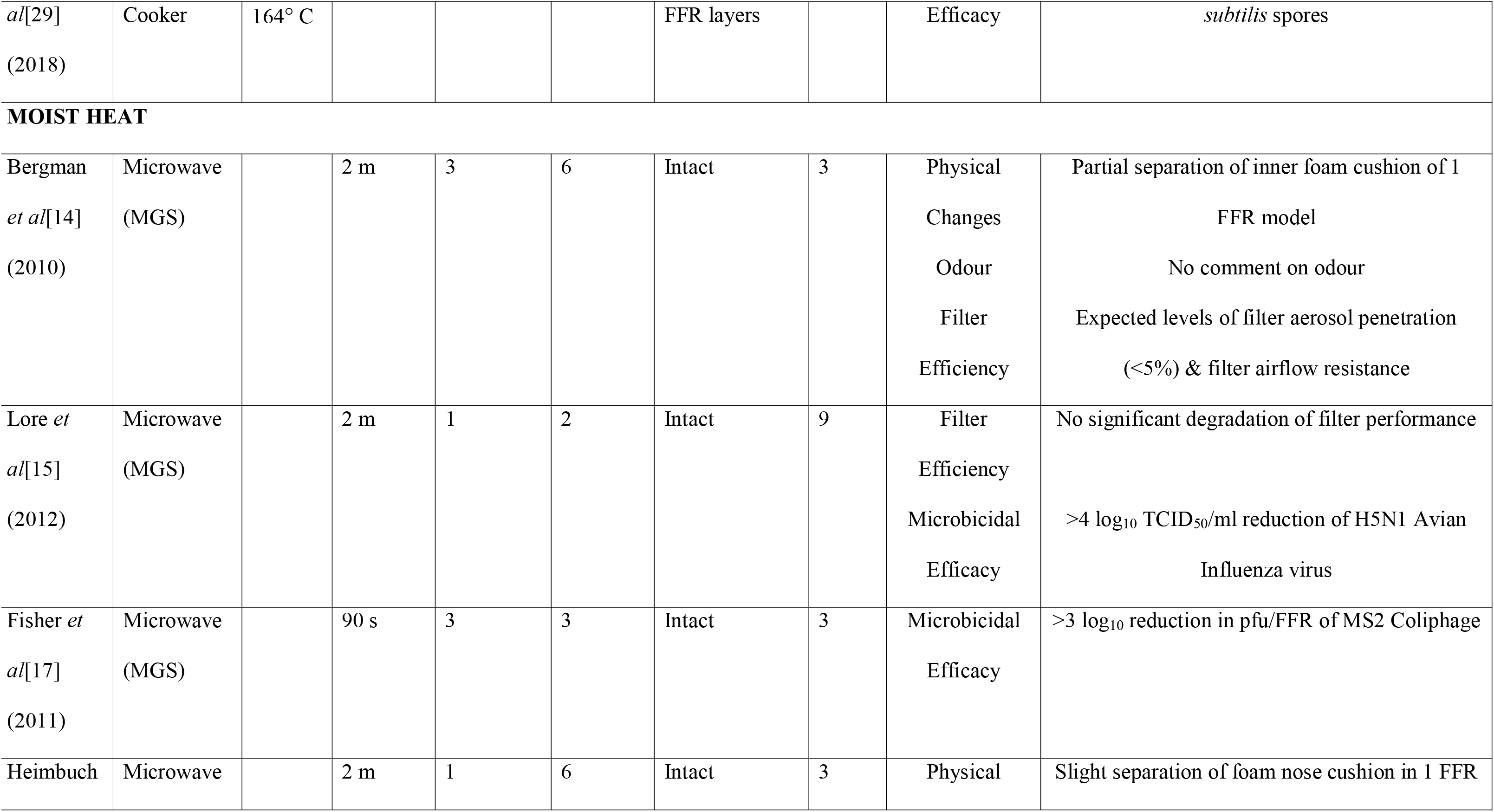
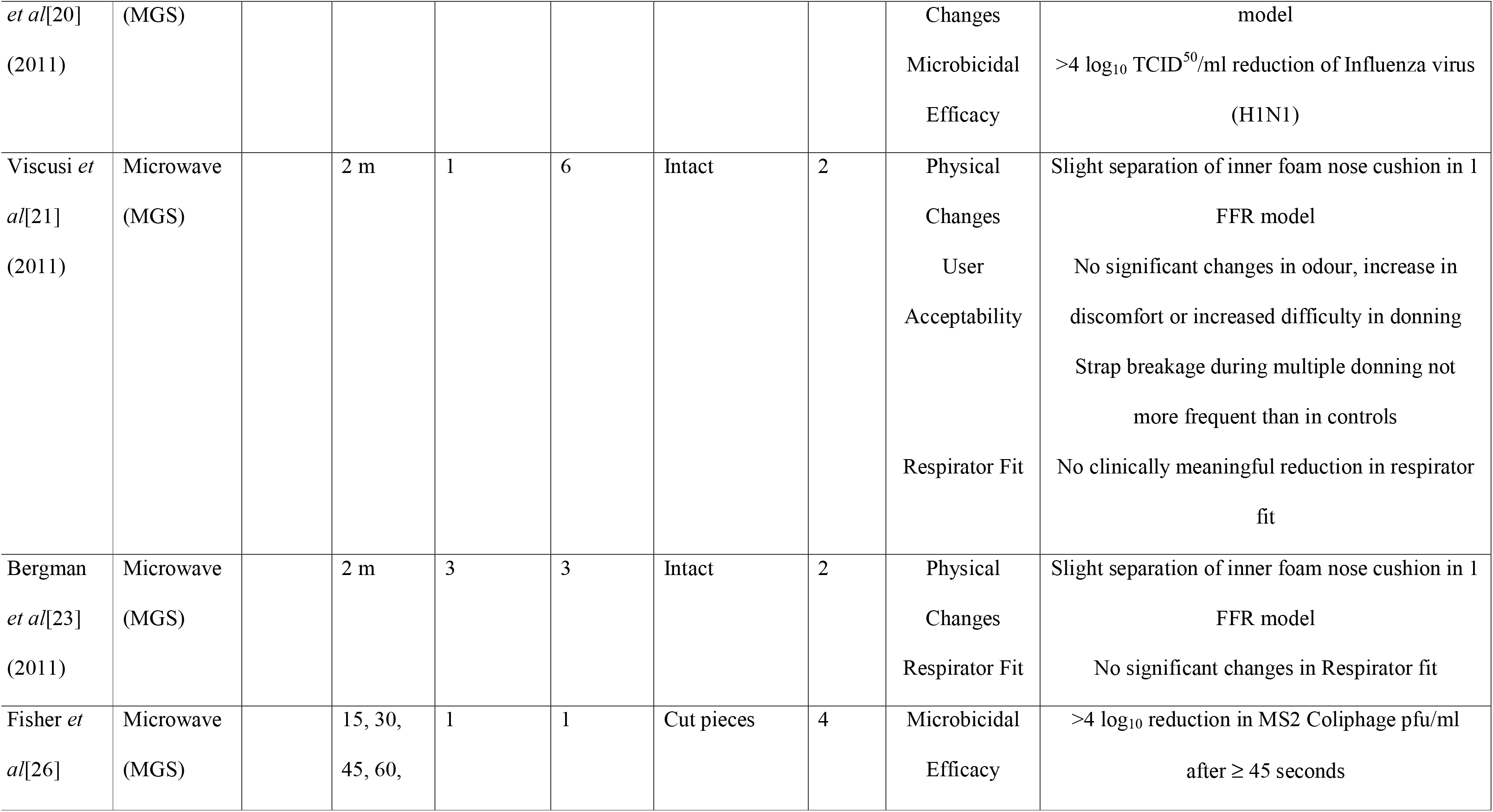
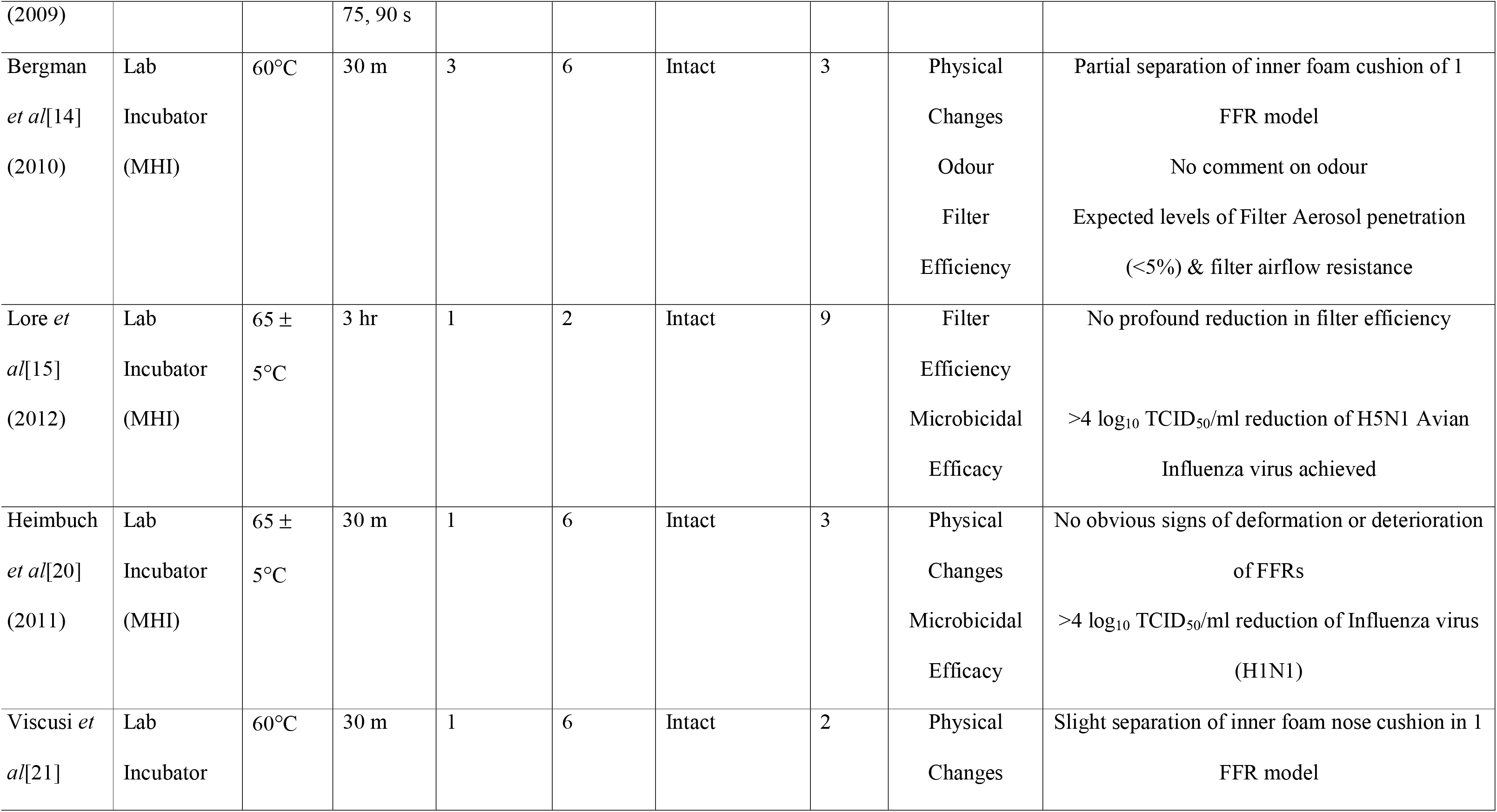
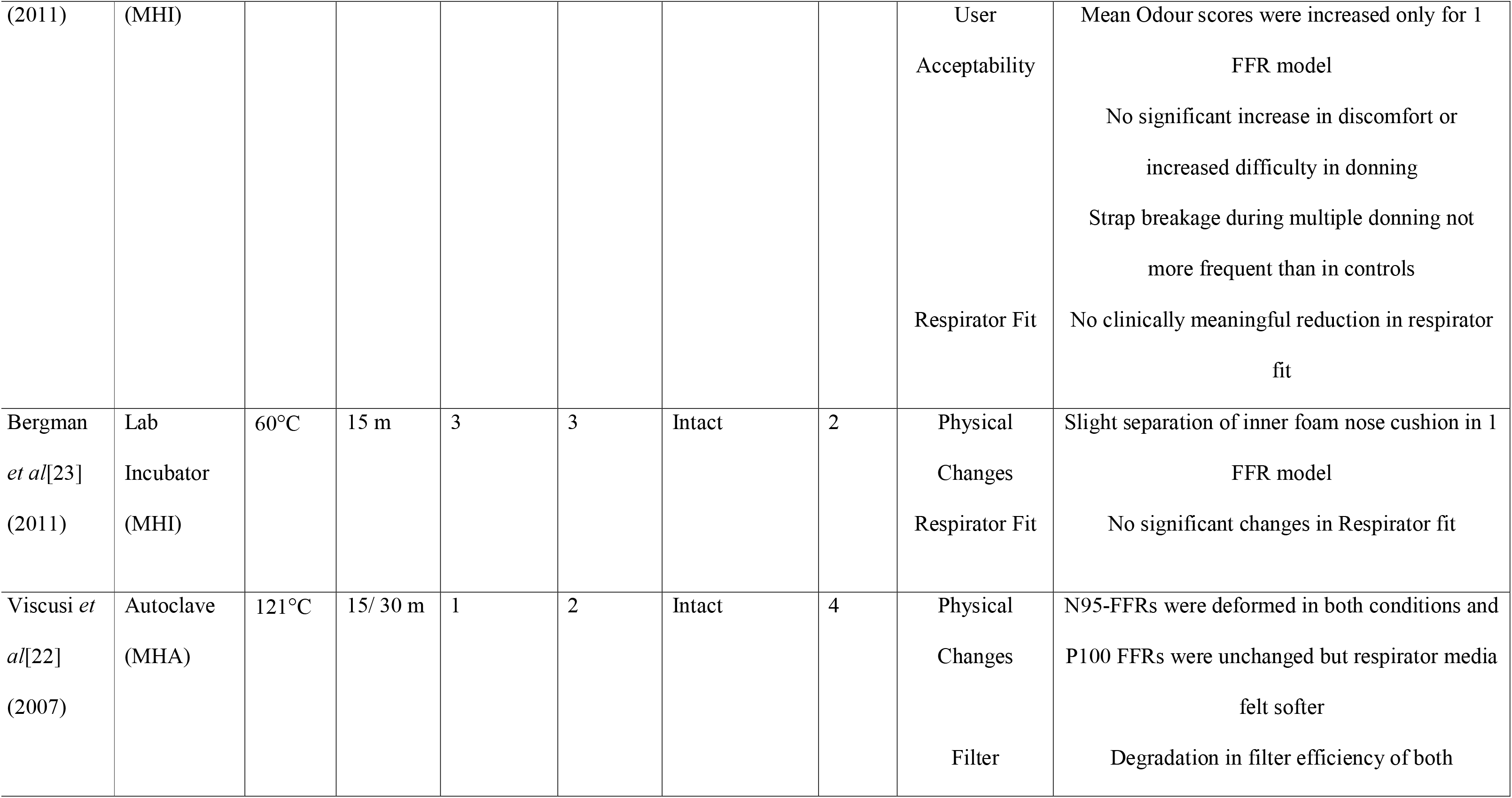
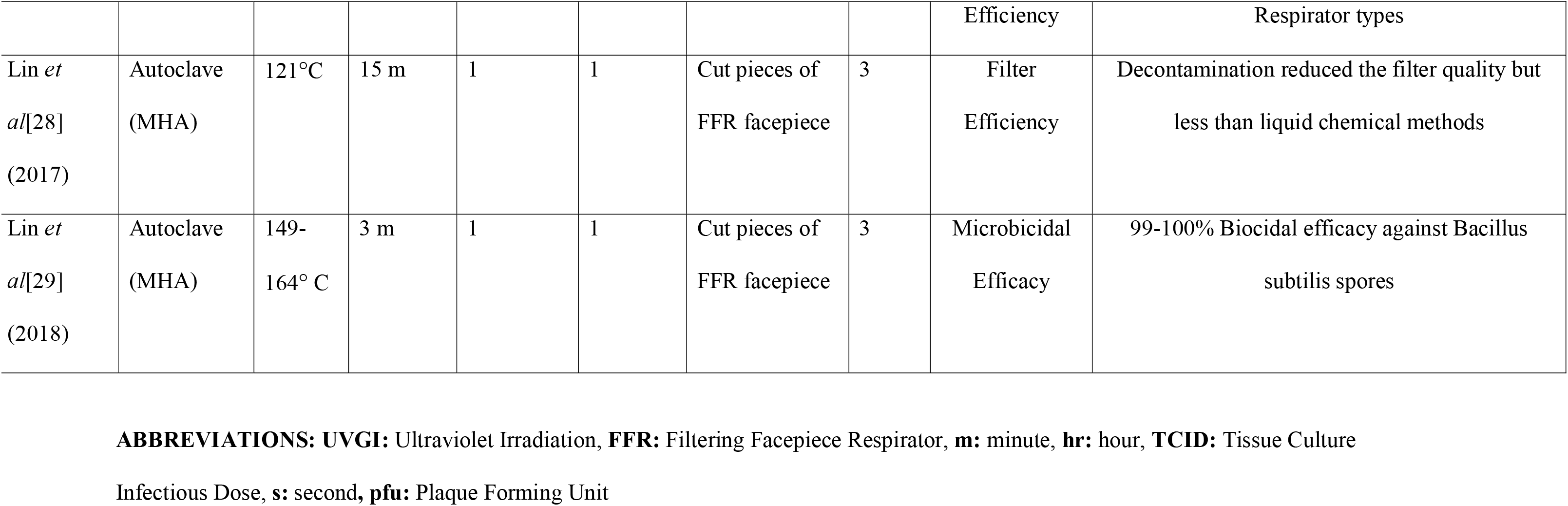
Summary of Characteristics of studies using Physical Decontamination methods, other than UVGI, for Reprocessing of FFRs.

##### iii. Dry Heat

Dry heat for reprocessing of FFRs has been evaluated in 4 studies[16,22,29,29] wherein microwave (MGI)[16,22,28,29], Hot Air Oven (DHO)[16,22] and Electric Rice Cooker (TERC)[28,29] have been used. Various parameters which have been evaluated against them are shown in Fig 2 and their exposure variables and results are summarized in Table 2. 3M8210 was the only known N95-FFR model which underwent reprocessing by any dry heat delivering method[28,29].

#### B. Gaseous Chemical Methods

Only 4 studies[14,16,22,27], prior to 2020, had evaluated a gaseous disinfection method for reprocessing of N95-FFRs. The methods used were Ethylene Oxide (EO)[14,16,22,27], Hydrogen peroxide in a Plasma Sterilizer (HPGP)[14,16,22,27] and Hydrogen Peroxide in vaporized form by using a commercial automated vapor generator[22]. Parameters against which they were evaluated; and their exposure variables and findings of the studies are provided in Fig 2 and Table 3, respectively. FFR models were not disclosed in any of the studies.

**Table 3:** Summary of Characteristics of studies using Gaseous Chemical Methods for Reprocessing of FFRs.

| Authors | Variables of Decontamination Methods |  |  |  | Variables of FFRs |  |  | Results |  |
| --- | --- | --- | --- | --- | --- | --- | --- | --- | --- |
|  | Disinfectant<br>Sterilizer | Packaging<br>Conditions | Duration | No. of<br>Deconta-<br>mination<br>Cycles | Total<br>no. of<br>Models<br>used | Part of<br>FFR<br>exposed | Repli-<br>cates | Parameters<br>Assessed | Summary of Results |
| Bergman<br>*et al*[14]<br>(2010) | Ethylene Oxide<br>Amsco®<br>Eagle® 3017 | Kept in<br>Tyvek®<br>pouches<br>6 FFR per<br>pouch | 1 hr<br>exposure<br>12 hr<br>aeration | 3 | 6 | Intact | 3 | Physical<br>Changes<br>Odour<br>Filter Efficiency | Partial separation of inner foam cushion<br>of 1 FFR model<br>No comment on odour<br>Expected levels of filter aerosol<br>penetration (<5%) & filter airflow<br>resistance |
| Viscusi *et*<br>*al*[16]<br>(2009) | Ethylene Oxide<br>3 M Steri-Vac<br>5XL | Individual<br>poly/paper<br>pouch | 1 hr<br>exposure<br>4 hr<br>aeration | 1 | 9<br>(6 N95<br>3 P100) | Intact | 3 | Physical<br>Changes<br>Filter Efficiency | No observable physical changes on<br>FFRs<br>Expected levels of filter aerosol<br>penetration (<5%) & filter airflow resistance |
| Viscusi *et al*[22]<br>(2007) | Ethylene Oxide<br>3 M Steri-Vac<br>4XL & 5 XL | Individual<br>poly/paper<br>pouch | 1 hr<br>exposure<br>4 hr<br>aeration | 1 | 2 | Intact | 4 | Physical<br>Changes<br>Filter Efficiency | Straps of P100 FFRs were slightly<br>darkened<br>Average penetration increased for both<br>respirator types but were within NIOSH<br>certification criteria |
| Salter *et al*[27]<br>(2010) | Ethylene Oxide<br>Amsco®<br>Eagle® 3017 | Individual<br>sterilization<br>pouch | 3 hr<br>exposure<br>12 hr<br>aeration | 1 | 6 | Intact | 3 | Presence of<br>Toxic Chemical<br>Residues | EO was not detected on any of the<br>model<br>Treated EO contained Diacetone<br>alcohol and a possible mutagen and<br>carcinogen, 2-hydroxyethyl acetate<br>(HEA) |
| Bergman<br>*et al*[14]<br>(2010) | H<sub>2</sub>O<sub>2</sub> Gas<br>Plasma (HPGP)<br>STERRAD® | Mylar/Tyvek<br>® pouch<br>6 samples per | 55 m cycle<br>time | 3 | 6 | Intact | 3 | Physical<br>Changes<br>Odour | No physical changes on FFRs<br>No comment on odour<br>25% (9/36) samples had aerosol |
|  | 100S | pouch |  |  |  |  |  | Filter Efficiency | penetration >5% suggestive of degradation in filter efficiency |
| Viscusi *et al*[16] (2009) | H<sub>2</sub>O<sub>2</sub> Gas<br>Plasma (HPGP)<br>STERRAD®<br>100S | Mylar/Tyvek<br>® pouch<br>6 samples per pouch | 55 m cycle<br>time | 1 | 9<br>(6 N95<br>3 P100) | Intact | 3 | Physical<br>Changes<br>Filter Efficiency | Metallic nose bands not as shiny as unexposed controls<br>Expected levels of Filter Aerosol penetration (<5%) & filter airflow resistance |
| Viscusi *et al*[22] (2007) | H<sub>2</sub>O<sub>2</sub> Gas<br>Plasma (HPGP)<br>STERRAD®<br>100S<br>STERRAD®<br>NX | Mylar/Tyvek<br>® pouch | 55 m<br>100 m | 1 | 2 | Intact | 4 | Physical<br>Changes<br>Filter Efficiency | Aluminium nosebands slightly tarnished with both cycles<br>Average penetration not significantly increased & remained within limit of NIOSH certification criteria for both respirator types and cycling conditions |
| Salter *et al*[27] (2010) | H<sub>2</sub>O<sub>2</sub> Gas<br>Plasma (HPGP)<br>STERRAD® | Sterilization<br>pouches | 55 m | 1 | 6 | Intact | 3 | Presence of<br>Toxic Chemical<br>Residues | No residues on FFRs<br>Sterilization cycle aborted when >6 FFRs were loaded in the sterilization |
|  | 100S |  |  |  |  |  |  |  | chamber |
| Bergman<br>*et al*[14]<br>(2010) | H<sub>2</sub>O<sub>2</sub> Vapor<br>(HPV)<br>Clarus® R HPV<br>Generator |  | 15 m dwell<br>125 m total<br>cycle time | 3 | 6 | Intact | 3 | Physical<br>Changes<br>Odour<br>Filter Efficiency | No physical changes on FFRs<br>No comment on odour<br>Expected levels of filter aerosol<br>penetration (<5%) & filter airflow<br>resistance |

**ABBREVIATIONS: FFR:** Filtering Facepiece Respirator, **hr:** Hour, **m:** Minute, **H<sub>2</sub>O<sub>2</sub>:** Hydrogen Peroxide,

#### C. Liquid Chemical Methods

Six different liquid decontamination methods have been evaluated on N95-FFRs in 8 studies[14,16,22,25-29]. These are Bleach[14,16,22,25-29], Liquid Hydrogen Peroxide (LHP)[14,22,27], Alcohols[22,28,29] including Ethanol and Isopropyl Alcohol, Mixed oxidants[27], Dimethyl Dioxirane[27] and Soap solution[22]. Parameters against which they were evaluated, their exposure variables and results of the studies are provided in Fig 2 and Table 4, respectively. Against Bleach, only known N95-FFR models evaluated were 3M8210 and Wilson SAF-T-FIT Plus (S4 Table). 3M8210 was the only known N95-FFR which was evaluated for Alcohols[28,29].

**Table 4:** Summary of Characteristics of Studies using Liquid & Miscellaneous Chemical Methods for Reprocessing of FFRs.

| Authors | Variables of Decontamination Methods | Variables of FFRs | Results |
| --- | --- | --- | --- |

|  | **Disinf-**<br>**ectant** | **Concen-**<br>**tration** | **Duration** | **No. of**<br>**Deconta-**<br>**mination**<br>**Cycles** | **Total**<br>**no. of**<br>**Models**<br>**used** | **Part of**<br>**FFR**<br>**exposed** | **Repli-**<br>**cates** | **Parameters**<br>**Assessed** | **Summary of Results** |
| --- | --- | --- | --- | --- | --- | --- | --- | --- | --- |
| Bergman<br>*et al*[14]<br>(2010) | Liquid<br>H<sub>2</sub>O<sub>2</sub> (LHP) | 6% | 30 m<br>Submersion | 3 | 6 | Intact | 3 | Physical Changes<br>Odour<br>Filter Efficiency | Staples were oxidized to varying degree<br>No comment on odour<br>Expected levels of Filter Aerosol penetration<br>(<5%) & filter airflow resistance |
| Viscusi *et al*[22]<br>(2007) | Liquid<br>H<sub>2</sub>O<sub>2</sub> (LHP) | 3%<br>6% | 30 m<br>submersion | 1 | 2<br>(1 N95<br>1 P100) | Intact | 4 | Physical Changes<br>Filter Efficiency | No observable changes on both respirator<br>types with 3% H<sub>2</sub>O<sub>2</sub> & slight fading of label<br>ink with 6% H<sub>2</sub>O<sub>2</sub><br>Average penetration within NIOSH<br>certification limit for both respirator types<br>& both concentrations |
| Salter *et al*[27] | Liquid<br>H<sub>2</sub>O<sub>2</sub> (LHP) | 3% | 30 m<br>submersion | 1 | 6 | Intact | 3 | Presence of Toxic<br>Chemical | No deposition of significant quantities of<br>toxic residues on FFRs |
| (2007) |  |  |  |  |  |  |  | Residues |  |
| Bergman<br>*et al*[14]<br>(2010) | NaOCl<br>(Bleach) | 0.6% | 30 m<br>Submersion | 3 | 6 | Intact | 3 | Physical Changes<br>Odour<br>Filter Efficiency | Metallic nosebands were tarnished, Staples<br>were oxidized to varying degree,<br>discoloured inner nose pads, dry to touch<br>All FFRs had a characteristic bleach odour<br>after overnight air drying<br>Expected levels of filter aerosol penetration<br>(<5%) & filter airflow resistance |
| Viscusi *et al*[16]<br>(2009) | NaOCl<br>(Bleach) | 0.6% | 30 m<br>Submersion | 1 | 9 | Intact | 3 | Physical Changes<br>Odour<br>Filter Efficiency | Metallic nose bands were tarnished<br>All FFRs had a scent of bleach and after<br>rehydration with water, increase in chlorine<br>off-gassing was measured<br>Expected levels of filter aerosol penetration<br>(<5%) & filter airflow resistance |
| Lin et al<sup>28</sup><br>(2017) | NaOCl<br>(Bleach) | 0.5% | 10 m<br>Submersion | 1 | 1 | Cut<br>pieces of facepiece | 3 | Filter Efficiency | Decontamination reduced the filter quality |
| Viscusi *et al*[22] (2007) | NaOCl (Bleach) | 0.52%<br>5.2% | 30 m<br>Submersion (both) | 1 | 2<br>(1 N95<br>1 P100) | Intact | 4 | Physical Changes<br>Filter Efficiency | Aluminium nose bands were tarnished at both concentrations<br>At 0.52% & 5.2% conc., average penetration for both respirator types were within NIOSH certification criteria |
| Lin *et al*[29] (2018) | NaOCl (Bleach) | 0.54%<br>2.7%<br>5.4% | NA<br>Inoculated | 1 | 1 | Cut<br>pieces of<br>Face-<br>piece | 3 | Microbicidal<br>Efficacy | 100% Biocidal efficacy against *Bacillus subtilis* spores at the lowest concentration |
| Vo *et al*[25] (2009) | NaOCl (Bleach) | 0.005/0.01/0.05/0.1/<br>0.25/0.5/<br>0.75% | 10 m<br>Submersion | 1 | 1 | Intact | 3 | Microbicidal<br>Efficacy | ≥0.5% bleach causes 4 log<sub>10</sub> reduction in pfu/ml of MS2 Coliphage |
| Fisher *et al*[26] | NaOCl (Bleach) | 0.0006%,<br>0.006%, | 2 m<br>Submersion | 1 | 1 | Cut<br>Coupons | 3 | Microbicidal<br>Efficacy | 0.6% bleach causes 4 log<sub>10</sub> reduction in pfu/ml of MS2 Coliphage |
| (2009) |  | 0.06%, 0.6% |  |  |  | of Face-<br>piece |  |  |  |
| Salter *et al*[27]<br>(2010) | NaOCl<br>(Bleach) | 0.6% | 30 m<br>Submersion | 1 | 6 | Intact | 3 | Physical changes<br>Odour<br>Presence of Toxic<br>Chemical<br>Residues | Corrosion of metal parts was noted<br>FFRs retained a bleach odour following an<br>off-gas period of 18 hour<br>Measured amount of residual chlorine was<br>below permissible exposure limit |
| Viscusi *et al*[22]<br>(2007) | Soap &<br>Water | 1g/L | 2 m<br>20 m<br>Submersion<br>(both) | 1 | 2<br>(1 N95<br>1 P100) | Intact | 4 | Physical Changes<br>Filter Efficiency | No physical changes observed for both<br>durations<br>Average penetration increased for both<br>durations and both respirators |
| Salter *et al*[27]<br>(2007) | Mixed<br>Oxidants | (10% Oxone,<br>6% Sodium<br>Chloride, 5%<br>Sodium | 30 m<br>submersion | 1 | 6 | Intact | 3 | Physical Changes<br>Odour<br>Presence of Toxic<br>Chemical | Oxidised metal parts<br>Left distinct odour on FFRs<br>No comment |
|  |  | Bicarbonate) |  |  |  |  |  | Residues |  |
| Salter *et al*[27]<br>(2007) | Dimethyl<br>Dioxirane | (10% Oxone,<br>10%<br>Acetone,<br>5% Sodium<br>Bicarbonate) | 30 m<br>submersion | 1 | 6 | Intact | 3 | Physical Changes<br>Odour<br>Presence of Toxic<br>Chemical<br>Residues | Oxidised metal parts<br>White residue accumulated on FFRs<br>Left distinct odour on FFRs<br>Retained in quantity by all 6 FFRs |
| **MISCELLANEOUS METHODS** |  |  |  |  |  |  |  |  |  |
| Heimbuch<br>*et al*[18] | NaOCl<br>(Bleach)<br>wipes | 0.9% | Surface<br>Cleaning of<br>outer and<br>inner layers | 3 | 3 | Intact | 3 | Microbicidal<br>Efficacy<br>Filter Efficiency<br>Mucin removal | 3-5 log reduction of *S. aureus* in the<br>presence of mucin<br>Mean particle penetration was <5%<br>No mucin detected, likely due to<br>interference in measurement assay by<br>NaOCl |
| Heimbuch<br>*et al*[18] | BAC wipes |  | Surface<br>Cleaning of<br>outer and | 3 | 3 | Intact | 3 | Microbicidal<br>Efficacy<br>Filter Efficiency | >4 log reduction of *S. aureus* in the presence<br>of mucin in most FFR samples<br>Mean particle penetration was <5% but |
|  |  |  | inner layers |  |  |  |  | Mucin removal | more than Bleach<br>Removal efficiency ranged from 21.47-76.41% but was poorer than inert wipes |
| Heimbuch<br>*et al*[18] | Inert wipes |  | Surface<br>Cleaning of<br>outer and<br>inner layers | 3 | 3 | Intact | 3 | Microbicidal<br>Efficacy<br>Filter Efficiency<br>Mucin Removal | No antibacterial activity<br>Mean particle penetration was <5%<br>Removal efficiency ranged from 21.47%-76.41% and better than BAC wipes |

**ABBREVIATIONS:** **FFR:** Filtering Facepiece Respirator, **H<sub>2</sub>O<sub>2</sub>:** Hydrogen Peroxide, **m:** Minute, **NaOCl:** Sodium Hypochlorite, **NIOSH:**
National Institute of Occupation Safety & Hygiene, **g/L:** Gram/Liter, ***S. aureus:*** *Staphylococcus aureus*, BAC: Benzalkonium Chloride

#### D. Miscellaneous Methods

In one study[18], commercial wipes of 0.9% Sodium Hypochlorite, Benzalkonium Chloride and an Inert material were evaluated for changes in filter efficiency and microbicidal efficacy by applying them on surface of N95-FFRs, as shown in Fig 2 & Table 4.

## Discussion

A pandemic of Influenza virus was always on the horizon and in 2009, it became reality. Researchers at NIOSH have been looking actively for finding a suitable method for their reprocessing since 2006 after the report of IOM Committee[11,22]. During 2007-2012, 12 studies evaluated reprocessing method for FFRs, most of them were conducted by or in collaboration with NIOSH[14-17,20-27]. In contrast, between 2013-2019, only 5 published studies had evaluated a reprocessing technique for N95-FFRs[19,20,28,29], with last study published by NIOSH in 2015[8]. Ongoing COVID-19 pandemic has brutally exposed the stalled progress in research to address this issue.

It has been shown that the surface stability of SARS-COV-2 on various surfaces lasts up to 3 days but this study didn’t include porous surfaces like that of respirators[37]. However, a study recently, showed it to be present on outer layer of surgical masks on day 7[38]. This recent data makes it imperative to decontaminate FFRs in between use as the risk of contact transmission without decontamination is considerable. Previously, CDC also discouraged reusing N95-FFRs whenever risk of contact transmission of a pathogen was high[6]. Furthermore, it is in larger global interest to find a suitable reprocessing method for N95-FFRs as they are not used frequently by HCWs in low to middle income countries (LMICs) while tackling pathogens against whom their use is mandatory such as Mycobacterium tuberculosis, as they are not available due to cost[39,40]. Finding a reprocessing method for FFRs will led to provision of adequate respiratory protection for HCWs in such resource limited settings.

A typical N95-FFR consists of facepiece covering mouth and nose, outer margin of which is lined to provide a face seal to the wearer. Two straps are attached to facepiece for fitting snugly at the back of head, a pliable metallic nose piece to facilitate bending at the nasal bridge and a foam cushion beneath it for the comfort of the wearer[41]. The function of an N95-FFR is to provide wearer an air supply free of particulates, including bioaerosols. This is facilitated by the main filtering layer of the respirator, also termed as Electret media, which is made up of non-woven, electrostatically charged polypropylene fibers which can capture the particulates in the incoming air[14]. However, the wearer must ensure that inhaled air should reach him through the facepiece and not through sides of the respirator. To ensure this, a wearer should undergo fit testing annually to determine the best respirator design and size for their facial features. Additionally, at the time of donning, wearer must ensure a user face-seal check to determine any air leak from the sides of respirator[41].

N95-FFRs are difficult to decontaminate owing to the porous nature of the main body and electrostatically charged nature of electret media. Any reprocessing or decontamination method, despite being microbiologically efficacious, shall be able to preserve the functioning of electret media, not physically affect various structural components compromising respirator fit and face-seal. Furthermore, due to proximity of N95-FFR to face, it should be devoid of harmful chemical residues, as they can be inhaled. Additional considerations for selecting a reprocessing method for reusing N95-FFRs for a healthcare facility are existing infrastructure, cost, turnaround time and throughput of the method[41]. Till date, whatever meagre research has been done, it has failed to find a reprocessing method which ticks all the boxes.

Physical (Energetic) methods such as application of moist heat, dry heat and irradiation have traditionally been the most commonly used methods for reprocessing healthcare items. Amongst them, Irradiation by UV-C (254 nm) rays (UVGI) has been the most frequently evaluated reprocessing methods for N95-FFRs, as shown in Fig 2 and Table 3. In all these studies, UVGI has shown to cause no damage to the physical appearance of FFRs, acceptable to users in terms of odor, donning ease and wear comfort, maintain respirator fit, preserve filter efficiency even after undergoing multiple cycles of decontamination and devoid of any toxic residues post-exposure.

Dose of irradiation is the most important variable for determining microbicidal efficacy of UVGI, which, in turn, is determined by irradiance at the surface of FFR and duration of exposure[19]. Total doses around 1-2 J/cm^2^ have shown to provide >4 log10 reduction of viruses inoculated on FFRs and around 5-6 J/cm^2^ against bacterial spores. Overall, UVGI has shown to be a suitable choice for reprocessing of FFRs, however, it is limited by varying exposure variables of UV dose used in multiple studies, as shown in Table 1. Furthermore, a study Fisher *et* al[24] concluded that the UV-C dose required for microbicidal efficacy is a function of the dose available to the electret medium, which in turn, is dependent on the penetrance (transmittance) of the layer above it. Hence, effective doses of UV-C for microbicidal efficacy will be model specific and needs to be established accordingly. We conclude that UVGI has great potential to be utilized as an effective decontamination method for N95-FFRs during this time of crisis however, more studies are needed to validate the various variables associated with the delivery of the UVGI method and respirator model specific doses will need to be established before it can be recommended.

Moist heat has been delivered in the form of steam generated in a microwave (MGS), benchtop lab incubator (MHI) at 60-70°C and in traditional Autoclave (MHA). Of them, MHA has shown to physically destroy FFRs and is deemed unsuitable for the purpose[22]. In MHI, exposure time has varied from 15-30 min and in MGS N95-FFRs are exposed for 90-120s. Multiple studies evaluating physical changes by both methods noticed partial separation of inner foam nose cushion. However, this was noticed for a particular FFR model (3M1870), where model identity was disclosed, but effect was not pronounced after undergoing multiple cycles of decontamination[14,23]. Both methods are shown to have no significant effect on user acceptability, respirator fit and filter efficiency till 3 cycles of decontamination[14,15,21,23]. More than 4 log_10_ reduction of enveloped viruses was demonstrated on N95-FFRs undergoing decontamination by MGS and MHI methods[15,17,20,26]. We are of opinion that these methods are low cost, easily doable in any setting, but require more validation in terms of other respirator models and cycles of decontamination, in future studies. MGS method is particularly suitable for implementation by individuals at home and smaller healthcare settings. Sparking due to placing metallic components in microwave has been a concern but it has not been noticed in MGS method[14].

Dry heat has been evaluated as a reprocessing method for FFRs in 4 studies[16,22,28,29] In two studies using DHO, FFRs were able to physically withstand temperatures till 80°C without affecting durability and filter efficiency[16,22] Electric rice cooker delivering temperature of 149-164°C has been used in studies from Taiwan[28,29] and one study from there found that exposure of an FFR model for 3 minutes was able to provide 99-100% biocidal efficacy against Bacillus subtilis spores[29]. In a study where microwave oven was used to deliver dry heat for 2 min, 2 of 9 respirator models were destroyed but in 7 models which withstood the treatment, filter efficiency was unchanged[16]. We opine that the literature is insufficient to either recommend or refute dry heat as a method of reprocessing for FFRs.

Exposure to Ethylene oxide (EO) and Hydrogen peroxide (H_2_O_2_) have been evaluated as a decontamination method for N95-FFRs simultaneously in 4 studies[14,16,22,27]. They are ideally suited for temperature sensitive articles hence, their use for reprocessing N95-FFRs is particularly promising. In these studies, FFRs have been exposed to EO and H_2_O_2_ (HPGP) in their respective sterilizers for standard cycling conditions. In addition, a study by Viscusi *et al*[22], vaporized H_2_O_2_ (HPV) generated in a commercial, automated vapor generator (BIOQUELL) was used for reprocessing of FFRs. Detailed cycling conditions of individual studies are given in Table 3. FFR models were not disclosed in any of the studies. After EO sterilization, FFRs didn’t showed any physical changes[14,16,22], or had offensive odor[14,16], and filter efficiency was also not degraded significantly[14,16,22] even after undergoing 3 cycles[14]. However, a study which focused on evaluating chemical residues post-exposure, found a possible carcinogen and mutagen, 2-hydroxyethyl acetate (HEA) on FFRs which had undergone EO sterilization[27]. No study yet has evaluated microbicidal efficacy of EO sterilization on FFRs though it is expected that this method will achieve adequate microbicidal efficacy. Overall, we opine that though EO has performed suitably in maintaining the physical architecture and filtration efficiency, increasing its safety profile by increasing aeration duration should be the topic of further research studies. Hence, it cannot be recommended at this point of time for reprocessing of N95-FFRs due to safety concerns.

Hydrogen peroxide provides microbicidal activity by way of generating free radicals and its degradation products are safe. In 3 studies, where HPGP was evaluated, no significant physical changes on the FFRs were noted[14,16,22]. Filter efficiency of 25% (9/36) respirators was noted to be degraded in one[14] of three[14,16,22] studies which evaluated. However, this effect was not noted when FFRs were treated with vaporized form[22,42]. In a commercial evaluation done for FDA by Batelle Institute on Clarus C HPV generator in 2016, no filter degradation was noted on 3M1870 even after undergoing 50 cycles of decontamination[42]. This system has been granted emergency use authorization (EUA) by FDA, after COVID-19 pandemic, for reprocessing N95-FFRs[43]. Concerns have been raised regarding throughput of HPGP as in a study authors noticed cycles were aborted in STERRAD® Sterilizer whenever >6 FFRs were placed[27]. This could be due to presence of cellulose in the straps of the respirators leading to absorption of H_2_O_2_P7]. No independent study prior to 2020 has evaluated microbicidal efficacy of H_2_O_2_ on FFRs though Batelle report showed 6 log reduction of *Geobacillus stearothermophilus* spores after undergoing reprocessing by HPV[42]. Overall, Hydrogen peroxide in gaseous form is a suitable option for reprocessing of N95-FFRs which needs to be evaluated rigorously for other parameters such as respirator fit and against other N95-FFRs. However, its availability is restricted to limited settings.

Submersion of FFRs in liquid disinfectants is a simple method of decontaminating them. Various liquid disinfectants which have been evaluated for this purpose namely liquid Hydrogen peroxide (LHP), household bleach and Alcohols. Hydrogen peroxide in 3% and 6% concentrations has been evaluated for decontamination of FFRs in 3 studies[14,22,27], of which two evaluated for physical changes and filter efficiency[14,22] whereas 1 study assessed for presence of toxic residues post-exposure[27]. Overall results were LHP oxidized staples of FFRs at 6% but not at 3% strength, filter efficiency of FFRs was maintained at both concentrations and they were devoid of toxic chemical residues after processing. Microbiological efficacy has not been studied yet in any study.

Bleach has been most frequently evaluated liquid disinfectant for reprocessing of FFRs. Overall bleach has been evaluated in 9 studies of which 1 used disinfectant wipes[18].

Exposure to bleach caused physical changes in the FFRs in terms of being stiff, mottled and tarnishing of metallic nosepieces[14,16,18,22]. Offensive odor from FFRs was noticed in most studies[14,16,27]. Furthermore, chlorine release has been noted when respirators were exposed to moisture, raising concerns regarding the safety of this method if a person breathes through it[16,27]. Though it has been found to have no significant degradation in the filter quality of the FFRs[14,16,18,22] and have excellent microbicidal efficacy[18,25,26,29], bleach is not safe to decontaminate FFRs. Alcohols (Ethanol and Isopropyl alcohol) have also been evaluated in 3 studies, but they are known to significantly degrade the filter efficiency due to removal of electrostatic charges from the electret media[22,28,29]. Hence, they don’t hold further merit in this discussion. Similar findings have been noted in one study which used soap & water for decontamination of FFRs[22].

A summary assessment of the body of literature on reprocessing of N95-FFRs has been provided in Fig 2. However, the findings of this systematic review and opinion of the authors should be assessed in light of limited literature available on this topic. Furthermore, readers should also consider the variability in exposure variables and methodological variabilities in the evaluated parameters within and between reprocessing methods. For example, to evaluate microbicidal efficacy, studies have used different micro-organisms and growth parameters accordingly while few included additional soiling challenges to mimic micro-organisms in human secretions. Few parameters have been evaluated only in few studies such as odor, wear comfort, and donning ease were evaluated objectively only in 1 study[21], respirator fit in 2 studies[21,23] and chemical safety in 1 study[27]. Hence, changes in these parameters which are not studied much, nevertheless are important, should be the focus of future studies. Furthermore, we didn’t do a meta-analysis as the studies were heterogeneous in terms of exposure variables and the number of studies conducted were less for a particular reprocessing method: parameter combination. As we write this review, a large body of literature on reprocessing of N95-FFRs has been already published till 30^th^ June 2020[44-58]. but when we did literature search, only few studies were published[44,45,49,58] and majority were in preprint, non-peer reviewed versions.

This systematic review is done to assess published literature, prior to COVID-19 pandemic, on reprocessing methods used to decontaminate N95-FFRs. This review may help administrators, infectious disease specialists and infection control personnel to formulate policies for effective utilization of single use, N95-FFRs to prevent respiratory transmission of SARS-COV-2. It will help researchers to find existing knowledge gaps in respirator reprocessing techniques and help them to design future studies. Furthermore, manufacturers may find it useful by knowing existing limitations and work their way around by developing new respirator material or design, more amenable to commonly available reprocessing techniques.

## Conclusions

To summarize, reusing N95-FFRs is need of the hour due to COVID-19 pandemic. Scientific progress was stalled after initial thrust provided by novel Influenza virus pandemic otherwise, current shortage of N95-FFRs for respiratory protection of HCWs would not have been of this humongous proportions. Consequently, HCWs have been forced to adopt measures which have little scientific support. Published literature is scant, but continuously updating on a daily basis, on studies evaluating reprocessing methods for N95-FFR. Besides microbiological efficacy, other factors such as physical changes, user acceptability of reprocessed FFRs, respirator fit, filter efficiency and chemical safety profile are of major consideration while selecting a reprocessing method for FFRs. Physical methods of decontamination are the most commonly evaluated methods for reprocessing of FFRs in the present scientific literature. However, except for UVGI, majority cause physical changes in the respirators in varying degrees. UVGI has varied widely in terms of dose of radiation delivered to FFRs and needs to be validated in more studies. Dose of UV-C irradiation which achieves satisfactory microbicidal efficacy needs to be determined specifically for each FFR model.

Use of low temperature sterilizing methods for reprocessing of N95-FFRs is promising. However, EO is not safe to use for reprocessing N95-FFRs whereas HPGP has been evaluated in very few studies yet and one study raised concerns about its effect on filter efficiency. Though emergency use approvals have been given to Hydrogen Peroxide STERRAD® Gas Plasma Sterilizer and BIOQUELL® Clarus C HPV generator, their presence is extremely limited worldwide, particularly in LMICs. Finding a suitable reprocessing method for N95-FFRs is also important from the perspective of infection control in LMICs. At present, promising technologies which need to be evaluated rigorously include UVGI and HPV. Other techniques such as MGS and MHI have shown to be efficacious against enveloped viruses and not compromise the filter efficiency up to 3 cycles of decontamination. Of them, MGS has an extremely short cycling duration and should be considered for emergency implementation in resource limited settings.

## Data Availability

All data available upon appropriate request

## Supporting Information

S1 Table: PRISMA Checklist

S2 Table: Search Strategy

S3 Table: Results of Quality Assessment & Risk Bias of Included Studies (After Inter-Author Agreement)

S4 Table: Summary of various reprocessing parameters evaluated for specific FFR models (where disclosed in included studies) by various reprocessing methods

**Abbreviations**
P: -Physical
O: -Odour
D: -Donning Ease & Wear Comfort
F: -Respirator Fit
E: Filter Efficiency
M: -Microbicidal Efficacy

**Respirator Manufacturers**
3M: 3M Company, Minneapolis MN
AP: Alpha Protech, Markham, Canada.
Cardinal: Cardinal Health, Inc, Dublin.
Gerson: Lois M Gerson Co, Inc, Middleboro, MA Inc,
KC: Kimberly Clark, Halyard Health Inc., Alpharaetta, GA.
Moldex: Moldex, Culver city, CA.
PA: Prestige Ameritech, North Richland Hills, TX.
Precept: Precept Medical products, Inc, Arden, NC.
Sperian: Honeywell Safety Products USA, Smithfield, RI.
Wilson: Wilson, Santa Ana, CA.
US Safety: Dentech Safety Specialists, Lenexa, KS.

